# Escape of SARS-CoV-2 501Y.V2 from neutralization by convalescent plasma

**DOI:** 10.1101/2021.01.26.21250224

**Authors:** Sandile Cele, Inbal Gazy, Laurelle Jackson, Shi-Hsia Hwa, Houriiyah Tegally, Gila Lustig, Jennifer Giandhari, Sureshnee Pillay, Eduan Wilkinson, Yeshnee Naidoo, Farina Karim, Yashica Ganga, Khadija Khan, Mallory Bernstein, Alejandro B. Balazs, Bernadett I. Gosnell, Willem Hanekom, Mahomed-Yunus S. Moosa, NGS-SA, COMMIT-KZN Team, Richard J. Lessells, Tulio de Oliveira, Alex Sigal

## Abstract

SARS-CoV-2 variants of concern (VOC) have arisen independently at multiple locations and may reduce efficacy of current vaccines targeted at the spike glycoprotein. We recently described the emergence of VOC in South Africa (501Y.V2 or PANGO lineage B.1.351) with mutations in the spike receptor-binding domain (RBD) and N-terminal domain (NTD). Here, using a live virus neutralization assay (LVNA), we compared neutralization of a first wave virus (B.1.1.117) versus the 501Y.V2 variant using plasma collected from adults hospitalized with COVID-19 from two South African infection waves, with the second wave dominated by 501Y.V2 infections. Sequencing demonstrated that infections in first wave plasma donors were with viruses harbouring none of the 501Y.V2-defining RBD or NTD mutations, except for one with E484K. 501Y.V2 virus was effectively neutralized by plasma from second wave infections and first wave virus was effectively neutralized by first wave plasma. In cross-neutralization, 501Y.V2 virus was poorly neutralized by first wave plasma, with an 8.4-fold drop in neutralization relative to first wave virus and a 15.1-fold drop relative to 501Y.V2 neutralization by second wave plasma. In contrast, second wave plasma neutralization of first wave virus was more effective, showing 4.1-fold decline relative to 501Y.V2 virus neutralization and 2.3-fold decline relative to first wave plasma neutralization. While we only tested one plasma elicited by E484K alone, this potently neutralized both variants. The observed effective neutralization of first wave virus by 501Y.V2 infection elicited plasma provides preliminary evidence that vaccines based on VOC sequences could retain activity against other circulating SARS-CoV-2 lineages.

---

Through genomic surveillance of the severe acute respiratory syndrome-related coronavirus 2 (SARS-CoV-2), a number of new variants have been identified with multiple mutations in the spike glycoprotein. We recently described the emergence of the 501Y.V2 (B.1.351) variant in South Africa, characterized by the K417N, E484K, and N501Y mutations in the spike receptor binding domain (RBD) as well as four substitutions and a deletion in the N-terminal domain (NTD) [1]. This variant was first detected in October 2020, and has rapidly become the dominant variant in South Africa with a frequency in January 2021 of 97% according to the GISAID (https://www.gisaid.org/hcov19-mutation-dashboard/).

The RBD is the main target of neutralizing antibodies (NAbs) elicited by SARS-CoV-2 infection, with the remaining activity directed at the NTD [2, 3]. All three amino acid residues in the RBD that carry mutations in 501Y.V2 interact directly with the human angiotensin-converting enzyme 2 (hACE2) receptor and form part of the epitopes for hACE2-blocking NAbs [4]. The E484 residue specifically is a hotspot for binding of highly potent NAbs [4]. In a number of separate *in vitro* studies using monoclonal antibodies (mAbs), mutations at E484 have emerged as immune escape mutations, often conferring broad cross-resistance to panels of mAbs [5, 6, 7, 8]. E484K also emerged during passage with convalescent plasma, leading to substantial drops in neutralization [9, 10]. Using a deep mutation scanning approach to determine the effect of individual mutations on neutralization by polyclonal sera, mutations at E484 were associated with the largest drops in neutralization [11].

South Africa experienced two SARS-CoV-2 infection waves to date (https://coronavirus.jhu.edu/map.html). The first wave peaked in July 2020 and consisted of viral variants which usually showed the D614G mutation but none of the defining mutations of 501Y.V2. These variants have been almost completely supplanted by 501Y.V2 variants in the second South African infection wave, peaking January 2021.

Coinciding with our initial report, there have been multiple studies showing that 501Y.V2 decreases neutralization capacity of polyclonal antibodies elicited by non-501Y.V2 SARS-CoV-2 infection or vaccination. The decrease ranges from relatively moderate [12, 13, 14, 15] to severe [16, 17, 18, 19, 20, 21]. Importantly, three clinical trials performed in South Africa during the second, 501Y.V2 infection wave reported dramatic decreases in vaccine efficacy. The Novavax NVX-CoV2373 subunit vaccine demonstrated a decrease in efficacy from 89.3% to 49.4% (https://ir.novavax.com/news-releases/news-release-details/novavax-covid-19-vaccine-demonstrates-893-efficacy-uk-phase-3). This trial also detected SARS-CoV-2 seroprevalence, and in the placebo arm there was no difference in infection frequency between participants who were seropositive for SARS-CoV-2 relative to those who were negative, indicating that previous infection with first wave, non-501Y.V2 virus does not protect against re-infection with 501Y.V2. The Johnson and Johnson adenovirus vectored single dose vaccine showed a reduced efficacy from 72% in the US to 57% in South Africa. (https://www.jnj.com/johnson-johnson-announces-single-shot-janssen-covid-19-vaccine-candidate-met-primary-endpoints-in-interim-analysis-of-its-phase-3-ensemble-trial). Most strikingly, the AstraZeneca ChAdOx1 AZD1222 chimpanzee adenoviral vectored vaccine showed only 10% efficacy against 501Y.V2 variants, compared to 75% efficacy against earlier variants circulating in South Africa [22]. The roll-out of this vaccine in South Africa is currently paused.

Here, using a live virus neutralization assay (LVNA), we measure the degree to which 501Y.V2 virus compromises neutralization elicited by natural infection with non-501Y.V2 variants. In addition, we measure the degree to which the earlier variants could escape the neutralizing response elicited to 501Y.V2 virus (Figure 1A). The possible relevance to vaccination is that an effective vaccine should be broadly protective against multiple variants, and this may depend on choice of variant sequence used in the design.

We used plasma samples from our ongoing longitudinal cohort that tracks COVID-19 cases enrolled at hospitals in Durban, South Africa [23]. We sampled participants weekly for the first month post-enrollment. At each timepoint a blood draw and combined nasopharyngeal/oropharyngeal swab was performed to obtain both the plasma and infecting virus. Swabs positive for SARS-CoV-2 were sequenced.

We chose plasma from 14 participants from the first South African infection wave where the infecting virus was successfully sequenced (Materials and Methods). Plasma samples were from blood drawn approximately 1 month post-symptom onset (Table S1), close to the antibody response peak [24]. Of the 14 participants, 13 did not show RBD or NTD mutations in the infecting virus. A single participant sampled in October 2020 showed the E484K escape mutation in the absence of the other 501Y.V2 mutations (Tables S2-S4). We had fewer participants from the second infection wave at the time of writing as most have not yet reached the 1 month post-symptom onset time-point for sampling. The second wave participants in this study were infected late December or early January 2021 (Figure 1B, Table S1). We were able to sequence three second wave participants where sequence allowed variant calling, two of which had good spike coverage (Figure 1B, Table S4). In all cases, the infecting variant was 501Y.V2. It is extremely likely that 501Y.V2 was also the infecting variant for the rest of the participants from infection wave 2, given the complete dominance of this variant in January 2021. For each second wave participant, our clinical team conducted a telephonic interview and examined clinical records to determine if the participant was also infected in the first South African infection wave. None of the participants showed evidence of being previously infected.

We outgrew first wave virus (Materials and Methods) from one participant during the first infection wave, and second wave, 501Y.V2 virus from a sample obtained in November 2020 through our genomic surveillance program (Figure 1B). We used a microneutralization live virus focus forming assay [25] which relies on a methylcellulose overlay to limit cell-free viral spread, resulting in a local infection focus. The focus is detected by an anti-SARS-CoV-2 spike antibody (Materials and Methods). We normalized the number of foci to the number of foci in the absence of plasma on the same plate to obtain the transmission index (Tx, [26]). This controls for experimental variability in the input virus dose between experiments. We mixed the virus with serially diluted plasma, then added the mixture to Vero E6 cells and counted infection foci after 28 hours using automated image analysis (Figure S1A, Figure 2A, Materials and Methods).

There was a clear reduction in neutralization capacity of 501Y.V2 by first wave plasma relative to neutralization of the homologous, first wave variant (Figure S1). 501Y.V2 also showed larger foci, likely reflecting a larger number of cells infected by one infected cell, or more rapid infection cycles (Figure 2A, Figure S1A). In order to compare foci of similar size we reduced the incubation time of 501Y.V2 infection to 18 hours. In order to detect some effect of first wave plasma on the 501Y.V2 variant, we tested more concentrated plasma (Figure 2A,B). To rule out infection saturation effects, we obtained a positive control monoclonal antibody with similar neutralization of first wave and 501Y.V2 variants. We then repeated the experiments (Figures S2-S4 show representative neutralization experiments for each participant plasma).

We observed the same trend in neutralization capacity as with the first set of experiments: there was a decline in the number of foci when first wave plasma was added to the homologous, first wave virus. This was strongly attenuated with 501Y.V2 (Figure 2B). When second wave, 501Y.V2 elicited plasma was used, it effectively neutralized the homologous, 501Y.V2 variant (Figure 2C). In contrast to the first wave plasma, the neutralization of the heterologous, first wave virus was clearly observed. Some of the foci were smaller at the higher antibody concentrations (Figure 2C, Figures S2-S4), possibly indicative of some reduction in cell-to-cell spread by neutralizing antibodies in the Vero E6 cell line.

The data from the focus forming assay at each dilution approximated a normal distribution (Figure S5) and we therefore used parametric statistics to describe it. We fitted the data for each participant to a sigmoidal function [27] with the dilution required to inhibit 50% of infection (*ID*_50_) as the only free parameter (Materials and Methods). For clarity, we plotted the data for each neutralization experiment as percent neutralization ((1−*Tx*)×100%, Materials and Methods, [16]), with neutralization represented by the 50% plaque reduction neutralization titer (*PRNT*_50_, [15]), the reciprocal of the *ID*_50_.

The A2051 monoclonal antibody was used as a positive control in each experiment (Figures S2-S4) and showed a similar neutralization response between variants (Figure 2D), indicating that focus number/size was not saturating. We also used a plasma pool from 3 study participants who did not have any indications of SARS-CoV-2 infection, and this plasma pool did not appreciably neutralize either variant (Figure 2D).

We then proceeded to quantify neutralization of the homologous virus and cross-neutralization. First wave virus infection was neutralized by first wave plasma, with some variability in neutralization capacity between first wave infected participants. It was also cross-neutralized by second wave, 501Y.V2 elicited plasma (Figure 2D). There was overlap between neutralization capacity of first wave and second wave plasma. In contrast, when the 510Y.V2 variant was used as the infecting virus, there was a clear separation between the neutralization capacity of the homologous second wave (Figure 2D) versus the heterologous first wave plasma. While the homologous plasma effectively neutralized 501Y.V2, the cross-neutralization mediated by first wave plasma was weaker, consistent with what is apparent when viewing the raw number of foci (Figure 2B-C, Figures S2-S4). Plasma elicited by the variant with the E484K mutation alone showed much stronger neutralization of both the first wave and 501Y.V2 virus relative any of the other plasma samples (Figure 2D).

The *PRNT*_50_ values showed a strong reduction in first wave plasma neutralization capacity of 501Y.V2 virus relative to the homologous first wave virus (Figure 2E). Excluding the plasma elicited by the virus with E484K mutation alone, which showed a very high *PRNT*_50_ for both variants, first wave plasma *PRNT*_50_ declined between 3.2 to 41.9-fold with the 501Y.V2 variant. In contrast, the decline in *PRNT*_50_ in cross-neutralization of first wave virus by second wave plasma was more attenuated. Here, the decline ranged between 1.6 to 7.2-fold relative to the homologous 501Y.V2 virus (Figure 2E).

Given the data approximated a normal distribution, we derived the mean neutralization across first wave (excluding the plasma elicited with the E484K only virus) and second wave participants (Figure 3). In both cases, neutralization showed a separation across all dilutions tested between the homologous and heterologous virus, where heterologous neutralization was lower. However, the separation was distinctly less for first wave virus neutralization by first wave versus second wave plasma (Figure 3). To quantify homologous versus heterologous neutralization capacity, we repeated the sigmoidal fit to the participant means and obtained the combined *PRNT*_50_. For first wave plasma neutralization of the homologous, first wave virus, *PRNT*_50_ was 344.0 with fit 95% confidence intervals of 275.4-458.0 (Figure 3 summary table, top left blue rectangle). For second wave plasma neutralization of the homologous, 501Y.V2 virus (Figure 3 summary table, bottom right blue rectangle), *PRNT*_50_ was 619.7 (517.8-771.5). Hence, 501Y.V2 elicits a robust antibody response in the participants tested. For cross-neutralization, first wave plasma neutralization of the heterologous, 501Y.V2 virus (Figure 3, bottom left yellow rectangle) was strongly attenuated across participants, with *PRNT*_50_ = 41.1. In contrast, second wave plasma neutralization of the heterologous, first wave virus (Figure 3, top right yellow rectangle) was more effective at *PRNT*_50_ = 149.7 (132.1-172.8). 95% confidence intervals did not overlap.

Fold-change decrease of first wave plasma neutralization of 501Y.V2 compared to homologous virus was 8.4. Fold-change decrease of second wave plasma neutralization of first wave compared to homologous virus was 4.1. However, absolute 501Y.V2 plasma neutralization capacity of first wave virus dropped only 2.3-fold compared to first wave plasma. In contrast, it decreased 15.1-fold when 501Y.V2 was cross-neutralized by first wave plasma (Figure 3).

The significance of these results is that 501Y.V2 is poorly neutralized by plasma elicited by non-501Y.V2 variants. However, 501Y.V2 infection elicited plasma not only effectively neutralized 501Y.V2, but also cross-neutralized the earlier variant within the observed range of the first wave plasma (Figure 2). This cross-neutralization is within the lower part of the neutralization capacity range elicited by the Pfizer BNT162b2 mRNA vaccine [12, 15, 13]. Due to potentially higher immunogenicity of 501Y.V2 virus according to the *PRNT*_50_, the plasma it elicits does not greatly under-perform the plasma elicited by earlier, non-501Y.V2 variants when neutralizing these earlier variants.

The larger focus size in 501Y.V2 relative to first wave virus is unlikely to influence these results. We performed 501Y.V2 infections with larger foci using the same infection incubation time as first wave virus, and also 501Y.V2 infections where focus size was similar using a shorter 501Y.V2 incubation time. The results showed similar trends. Furthermore, neutralization by the monoclonal antibody control indicated that the LVNA system could effectively read out neutralization for both variants (Figure 2D, Figure S2). 501Y.V2 variants vary in some of their mutations. The variant we used has an L18F mutation in the NTD which currently occurs in about a quarter of 501Y.V2 variants (GISAID). Other current and future 501Y.V2 variants can be examined to track changes in neutralization and cross-neutralization. An important question in the interpretation of the results is whether the second wave infected participants were also infected in the first infection wave. Our clinical team conducted telephonic interviews and investigated the clinical charts and found no evidence of previous SARS-CoV-2 infection. While previous infection may still be missed despite these measures, we it is unlikely to have occurred in all the second wave participants, yet the neutralization response between the participants was very similar (Figure 2). Lastly, while we and others in the field measured plasma neutralization, how well this correlates to protection at the mucosal surface where initial infection takes place is yet unclear.

The plasma elicited by virus with the E484K mutation alone showed the strongest neutralization both of the first wave and the 501Y.V2 virus relative to any of the other plasma samples tested (Figure 2). Because we only found one participant in this category, this result is difficult to interpret: it may be due to high immunogenicity or because of participant specific factors. Our clinical data does not show prolonged SARS-CoV-2 shedding in this participant or other unusual features (Table S1). This result highlights the importance of sequencing the infecting virus, and requires further investigation.

The recent Novavax, Johnson and Johnson, and AstraZeneca South African vaccine trial results indicate that the 501Y.V2 variant may lead to a decrease in vaccine efficacy. The loss of neutralization capacity in 501Y.V2 infection we quantified among the vaccinated participants in the AstraZeneca trial [22] shows that loss of neutralization may be associated with loss of vaccine efficacy. Loss of vaccine efficacy may also be mediated by escape from T cell immunity, although we believe this is less likely due to the diversity of HLA alleles in the population, which may curtail the ability of an escape variant which evolved in one individual to escape T cell immunity in another. If loss of vaccine efficacy proves to require vaccine redesign, the results presented here may be the first indication that a vaccine designed to target 501Y.V2 may also effectively target other SARS-CoV-2 variants.

**Figure 1:**
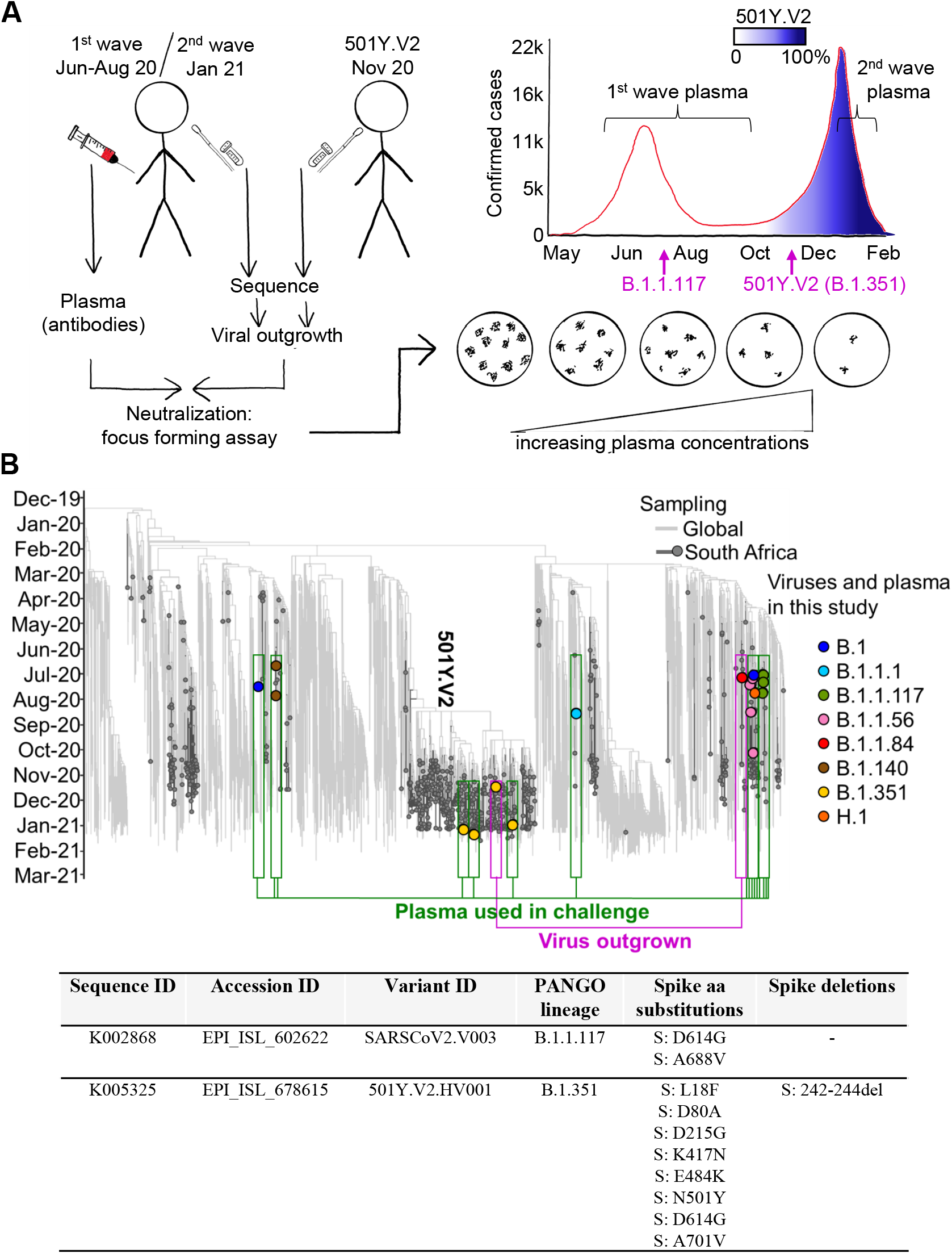
Study design and sequences of SARS-CoV-2 variants. (A) We obtained convalescent plasma and sequenced the matching infecting virus in the first and second SARS-CoV-2 infection waves in South Africa. A first wave variant lacking the RBD and NTD mutations characterizing 501Y.V2 was outgrown from one participant infected in the first South African infection wave, and 501Y.V2 was outgrown from a participant at the beginning of the second wave. Neutralization performed was by i) first wave plasma of first wave virus; ii) second wave plasma of 501Y.V2 virus; iii) first wave plasma of 501Y.V2 virus; iv) second wave plasma of first wave virus. A focus forming microneutralization assay was used to quantify neutralization. (B) Phylogenetic relationships and mutations in virus sequences. Variants which elicited the antibody immunity in the plasma samples are highlighted in green boxes. Variants which were outgrown are highlighted in magenta boxes. Y-axis denotes time of sampling. Table shows mutations present in spike for the SARS-CoV-2 variants used in the LVNA. See Tables S2-S4 for a complete list of mutations in the viral genomes of both the variants used in LVNA, and the sequenced variants eliciting the plasma immunity.

**Figure 2:**
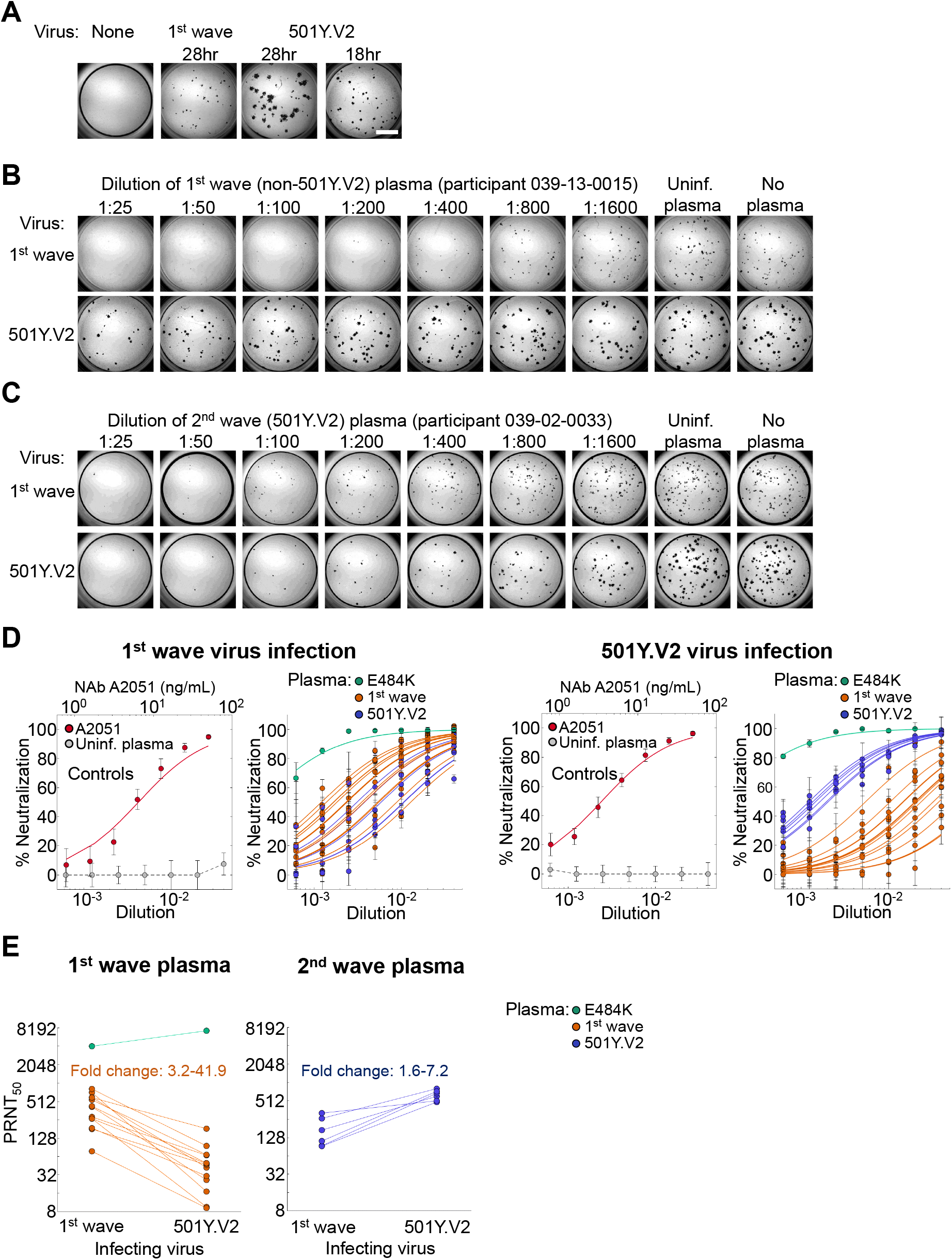
Neutralization of first infection wave and 501Y.V2 variants by convalescent plasma from South African first and second wave infections. (A) Focus formation in the absence of plasma when infection is by a first wave versus the 501Y.V2 viral variant. Incubation time for 501Y.V2 focus formation was reduced to 18 hours to obtain similar focus size. Scale bar is 2mm. (B) A representative focus forming assay using plasma from first wave infected participant 039-13-0015. First row is neutralization of infection by first wave virus, second row is neutralization of 501Y.V2. Columns are plasma dilutions, ranging from 1:25 to 1:1600. Last two columns are plasma from a pool of uninfected participants and the no plasma control. (C) Representative neutralization using plasma from second wave infected participant 039-02-0033. (D) Left two plots represent controls and plasma neutralization when infection is with first wave virus and right two plots when infection is with 501Y.V2. Points are means and standard errors of percent neutralization from 3 or 4 independent experiments for convalescent plasma from first wave (n=14) or second wave (n=6) participants, or 10 independent experiments for the controls. Solid lines of the corresponding colour are fitted values per participant using a fit to a sigmoidal equation. First plot is neutralization of first wave virus by neutralizing antibody A2051. *PRNT*_50_ = 6.5 ng/mL (3.9-9.1 ng/mL). Neutralization by plasma from uninfected participants is represented by the grey points. Second plot is neutralization of first wave virus by plasma from convalescent participants. Groups are first infection wave are (red), second wave (blue), and virus with E484K only (green). Third plot is the control neutralization of the 501Y.V2 variant by A2051. *PRNT*_50_ = 3.5 ng/mL (2.9-4.1 ng/mL).Forth plot is neutralization of 501Y.V2 virus by plasma from convalescent participants. Groups are first infection wave are (red), second wave (blue), and virus with E484K only (green). (E) Decline in *PRNT*_50_ in cross-neutralization of heterologous virus. Left plot is first wave plasma neutralization of first wave versus 501Y.V2 virus, and right plot is second wave plasma neutralization of 501Y.V2 versus first wave virus. For first wave plasma fold-change in *PRNT*_50_ was calculated as *PRNT*_50_ of first wave divided by *PRNT*_50_ of 501Y.V2. For second wave plasma, fold change was calculated as *PRNT*_50_ of 501Y.V2 divided by *PRNT*_50_ of first wave virus. Fold change ranged from 3.2 to 41.9 for first wave plasma, and 1.6 to 7.2 for second wave plasma. For first wave plasma, *PRNT*_50_ of plasma elicited by E484K virus was excluded.

**Figure 3:**
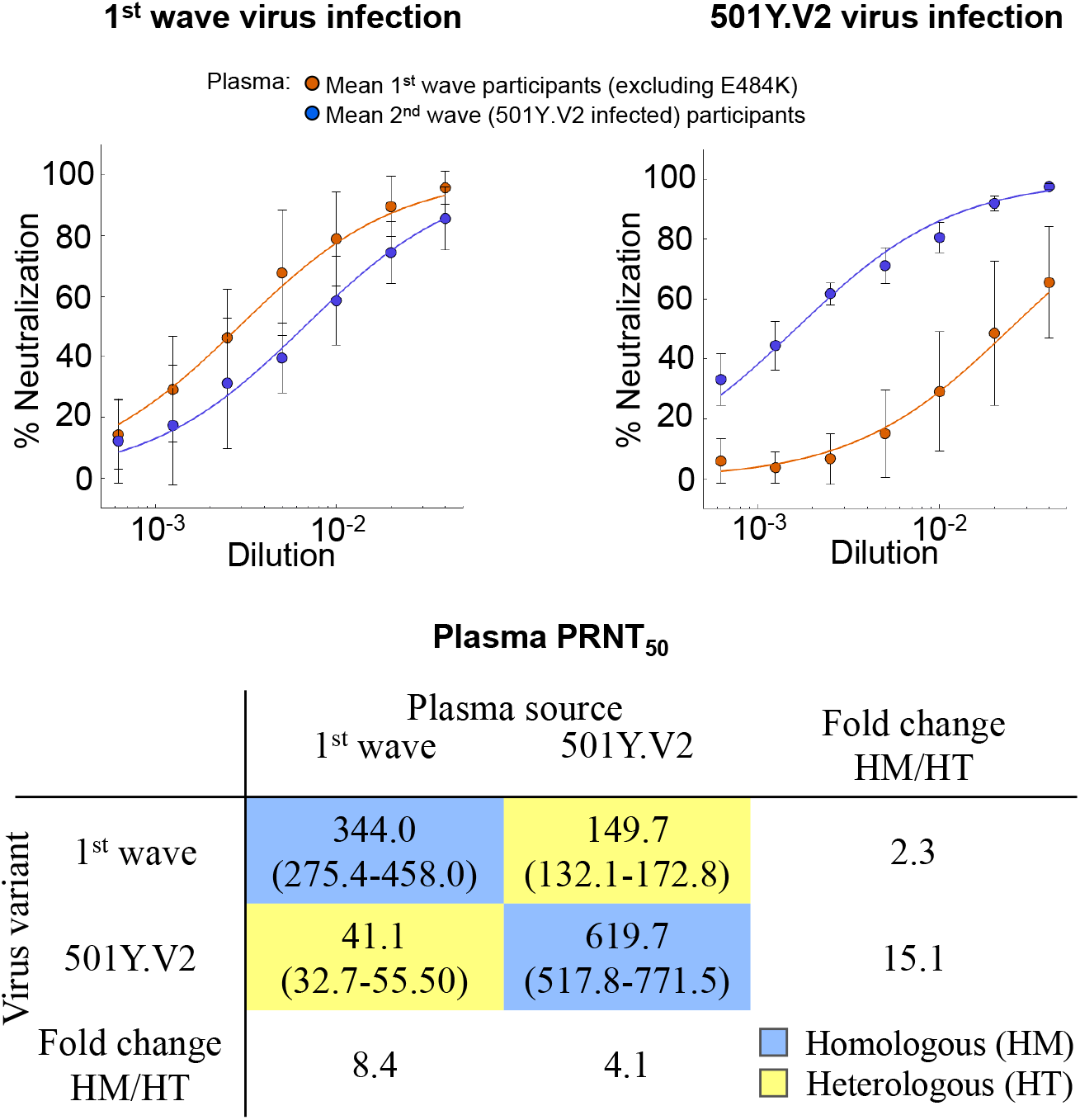
Cross-neutralization of first infection wave and 501Y.V2 virus across all participants from two infection waves. Left plot shows neutralization of infection by first wave virus, and right plot shows neutralization of 501Y.V2. Sigmoidal fits were performed to the means of first (red points) and second wave (blue points) plasma neutralization results across all participants excluding the participant with plasma immunity elicited by the viral variant containing the E484K mutation alone. Shown are means and standard deviations of n=13 first wave infected plasma donors and n=6 second wave infected plasma donors. Summary table shows plasma *PRNT*_50_ as a function of plasma source (columns) and infecting viral variant (rows). Blue rectangles highlight homologous neutralization where virus and infection wave are matched, and yellow rectangles highlight cross-neutralization where virus and plasma are from different infection waves.

## Material and methods

### Ethical statement

Nasopharyngeal/oropharyngeal swab samples and plasma samples were obtained from six hospitalized adults with PCR-confirmed SARS-CoV-2 infection enrolled in a prospective cohort study approved by the Biomedical Research Ethics Committee (BREC) at the University of KwaZulu-Natal (reference BREC/00001275/2020). The 501Y.V2 variants were obtained from residual nasopharyngeal/oropharyngeal samples used for routine SARS-CoV-2 diagnostic testing by the National Health Laboratory Service, through our SARS-CoV-2 genomic surveillance program (BREC approval reference BREC/00001510/2020).

### Whole genome sequencing, genome assembly and phylogenetic analysis

cDNA synthesis was performed on the extracted RNA using random primers followed by gene specific multiplex PCR using the ARTIC V3 protocol. Briefly, extracted RNA was converted to cDNA using the Superscript IV First Strand synthesis system (Life Technologies, Carlsbad, CA) and random hexamer primers. SARS-CoV-2 whole genome amplification was performed by multiplex PCR using primers designed on Primal Scheme (http://primal.zibraproject.org/) to generate 400bp amplicons with an overlap of 70bp that covers the 30Kb SARS-CoV-2 genome. PCR products were cleaned up using AmpureXP purification beads (Beckman Coulter, High Wycombe, UK) and quantified using the Qubit dsDNA High Sensitivity assay on the Qubit 4.0 instrument (Life Technologies Carlsbad, CA). We then used the Illumina® Nextera Flex DNA Library Prep kit according to the manufacturer’s protocol to prepare indexed paired end libraries of genomic DNA. Sequencing libraries were normalized to 4nM, pooled and denatured with 0.2N sodium acetate. 12pM sample library was spiked with 1% PhiX (PhiX Control v3 adapter-ligated library used as a control). We sequenced libraries on a 500-cycle v2 MiSeq Reagent Kit on the Illumina MiSeq instrument (Illumina, San Diego, CA). We assembled paired-end fastq reads using Genome Detective 1.126 (https://www.genomedetective.com) and the Coronavirus Typing Tool. We polished the initial assembly obtained from Genome Detective by aligning mapped reads to the references and filtering out low-quality mutations using the bcftools 1.7-2 mpileup method. Mutations were confirmed visually with bam files using Geneious software (Biomatters Ltd, Auckland, New Zealand). All of the sequences were deposited in GISAID (https://www.gisaid.org/). We retrieved all South African SARS-CoV-2 genotypes from the GISAID database as of 11 January 2021 (N=2704). We initially analyzed South African genotypes against the global reference dataset (N=2592) using a custom pipeline based on a local version of NextStrain. The pipeline contains several python scripts that manage the analysis workflow. It performs alignment of genotypes in MAFFT, phylogenetic tree inference in IQ-Tree20, tree dating and ancestral state construction and annotation (https://github.com/nextstrain/ncov).

### Cells

Vero E6 cells (ATCC CRL-1586, obtained from Cellonex) were propagated in complete DMEM with 10% fetal bovine serum (Hylone) containing 1% each of HEPES, sodium pyruvate, L-glutamine, and nonessential amino acids (Sigma-Aldrich). Cells were passaged every 3-4 days. H1299 cells were propagated in complete RPMI with 10% fetal bovine serum containing 1% each of HEPES, sodium pyruvate, L-glutamine, and non-essential amino acids and and passaged every second day.

### H1299-E3 cell line for first passage SARS-CoV-2 outgrowth

The H1299-H2AZ clone with nuclear labelled YFP was constructed to overexpress ACE2 as follows: VSVG-pseudotyped lentivirus containing the human ACE2 was generated by co-transfecting 293T cells with the pHAGE2-EF1alnt-ACE2-WT plasmid along with the lentiviral helper plasmids HDM-VSVG, HDM-Hgpm2, HDM-tat1b and pRC-CMV-Rev1b using TransIT-LT1 (Mirus) transfection reagent. Supernatant containing the lentivirus was harvested two days after infection, filtered through a 0.45*µ*m filter (Corning) and used to spinfect H1299-H2AZ at 1000 rcf for 2 hours at room temperature in the presence of 5 *µ*g/mL polybrene (Sigma-Aldrich). ACE-2 transduced H1299-H2AZ cells were then subcloned at the single cell density in 96-well plates (Eppendorf) in conditioned media derived from confluent cells. After 3 weeks, wells were trypsinized (Sigma-Aldrich) and plated in two replicate plates, where the first plate was used to determine infectivity and the second was stock. The first plate was screened for the fraction of mCherry positive cells per cell clone upon infection with SARS-CoV-2 mCherry expressing spike pseudotyped lentiviral vector 1610-pHAGE2/EF1a Int-mCherry3-W produced by transfecting as above. Screening was performed using a Metamorph-controlled (Molecular Devices, Sunnyvale, CA) Nikon TiE motorized microscope (Nikon Corporation, Tokyo, Japan) with a 20x, 0.75 NA phase objective, 561 laser line, and 607 nm emission filter (Semrock, Rochester, NY). Images were captured using an 888 EMCCD camera (Andor). Temperature (37°C), humidity and CO2 (5%) were controlled using an environmental chamber (OKO Labs, Naples, Italy). The clone with the highest fraction of mCherry expression was expanded from the stock plate and denoted H1299-E3. This clone was used in the outgrowth.

### Viral Outgrowth

All live virus work was performed in Biosafety level 3 containment using Africa Health Research Institute biosafety committee approved protocols for SARS-CoV-2. For first wave virus, a T25 flask (Corning) was seeded with Vero E6 cells at 2 × 10^5^ cells/ml and incubated for 18-20 hours. After 1 DPBS wash, the sub-confluent cell monolayer was inoculated with 500*µ*L universal transport medium (UTM) diluted 1:1 with growth medium and filtered through a 0.45*µ*M filter. Cells were incubated for 1 hour. Flask was then filled with 7mL of complete growth medium and checked daily for cytopathic effect (CPE). Four days post infection, supernatants of the infected culture were collected, centrifuged at 300 rcf for 3 minutes to remove cell debris, and filtered using a 0.45*µ*M filter. Viral supernatant was aliquoted and stored at −80°C. For 501Y.V2 variants, we used H1299-ACE2-E3 cells for initial isolation followed by passage into Vero E6 cells. H1299-ACE2-E3 cells were seeded at 1.5 × 10^5^ cells/ml and incubated for 18-20 hours. After 1 DPBS wash, the sub-confluent cell monolayer was inoculated with 500*µ*L universal transport medium (UTM) diluted 1:1 with growth medium and filtered through a 0.45*µ*M filter. Cells were incubated for 1 hour. Wells were then filled with 3mL of complete growth medium. 8 days postinfection, cells were trypsinized, centrifuged at 300 rcf for 3 minutes and resuspended in 4mL growth medium. 1mL was added to Vero E6 cells that had been seeded at t 2 × 10^5^ cells/ml 18-20 hours earlier in a T25 flask (approximately 1:8 donor-to-target cell dilution ratio) for cell-to-cell infection. Coculture of H1299-ACE2-E3 and Vero E6 cells was incubated for 1 hour and flask was then filled with 7mL of complete growth medium and incubated for 6 days. Viral supernatant was aliquoted and stored at −80°C or further passaged in Vero E6 cells as above. Two isolates were outgrown, 501Y.V2.HV001 and 501Y.V2.HVdF002. The second isolate showed fixation of furin cleavage site mutations during outgrowth in Vero E6 cells and was not used except for data presented in Figure S1.

### Microneutralization using focus forming assay

For plasma from first wave donors, we first quantified spike RBD IgG by enzyme-linked immunosorbent assay (ELISA) using monoclonal antibody CR3022 as a quantitative standard, (n = 13 excluding participant 039-13-0103 for which ELISA data was not available). The mean concentration was 23.7 *µ*g/mL *pm* 19.1 *µ*g/mL, (range 5.7 - 62.6 *µ*g/mL). In comparison, uninfected donor controls had a mean of 1.85 *µ*g/mL *pm* 0.645 *µ*g/mL. To quantify neutralization, Vero E6 cells were plated in an 96-well plate (Eppendorf or Corning) at 30,000 cells per well 1 day pre-infection. Importantly, before infection approximately 5ml of sterile water was added between wells to prevent more rapid drying of wells at the edge which we have observed to cause edge effects (lower number of foci). Plasma was separated from EDTA-anticoagulated blood by centrifugation at 500 rcf for 10 minutes and stored at −80°C. Aliquots of plasma samples were heat-inactivated at 56°C for 30 minutes, and clarified by centrifugation at 10,000 rcf for 5 minutes, where the clear middle layer was used for experiments. Inactivated plasma was stored in single use aliquots to prevent freeze-thaw cycles. For experiments, plasma was serially diluted two-fold from 1:100 to 1:1600, where this is the concentration during the virus-plasma incubation step before addition to cells and during the adsorption step. As a positive control, the GenScript A02051 anti-spike mAb was added at concentrations listed in the figures. Virus stocks were used at approximately 50 focus-forming units (FFU) per microwell and added to diluted plasma; antibody-virus mixtures were incubated for 1 hour at 37°C, 5% CO2. Cells were infected with 100*µ*L of the virus-antibody mixtures for one hour, to allow adsorption of virus. Subsequently, 100*µ*L of a 1x RPMI 1640 (Sigma-Aldrich R6504), % carboxymethylcellulose (Sigma-Aldrich C4888) overlay was added to the wells without removing the inoculum. Cells were fixed at 28 hours post-infection using 4% paraformaldehyde (Sigma-Aldrich) for 20 minutes. For staining of foci, a rabbit anti-spike monoclonal antibody (mAb BS-R2B12, GenScript A02058) was used at 0.5*µ*g/mL as the primary detection antibody. Antibody was resuspended in a permiabilization buffer containing 0.1% saponin (Sigma-Aldrich), 0.1% BSA (Sigma-Aldrich), and 0.05% tween (Sigma-Aldrich) in PBS. Plates were incubated with primary antibody overnight at 4°C, then washed with wash buffer containing 0.05% tween in PBS. Secondary goat anti-rabbit horseradish peroxidase (Abcam ab205718) was added at 1 *µ*g/mL and incubated for 2 hours at room temperature with shaking. The TrueBlue peroxidase substrate (SeraCare 5510-0030) was then added at 50*µ*L per well and incubated for 20 minutes at room temperature. Plates were then dried for 2 hours and imaged using a Metamorph-controlled Nikon TiE motorized microscope with a 2x objective. Automated image analysis was performed using a Matlab2019b (Mathworks) custom script, where focus detection was automated and did not involve user curation. Image segmentation steps were stretching the image from minimum to maximum intensity, local Laplacian filtering, image complementation, thresholding and binarization. Two plasma donors initially measured from the South African second infection wave did not have detectable neutralization of either 501Y.V2 or the first wave variant and were not used in the study.

### Statistics and fitting

All statistics and fitting were performed using Matlab2019b. Neutralization data was fit to

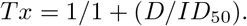

Here Tx is the number of foci normalized to the number of foci in the absence of plasma on the same plate at dilution D. To visiualize the data, we used percent neutralization, calculated as (1-Tx) ×100%. Negative values (Tx¿1, enhancement) was represented as 0% neutralization. Fit to a normal distribution used Matlab2019b function normplot, which compared the distribution of the Tx data to the normal distribution (see https://www.mathworks.com/help/stats/normplot.html).

## Data Availability

All sequence data is available at GISAID.

## Acknowledgements

This work was supported by the Bill and Melinda Gates Investment INV-018944 (AS) and by the South African Medical Research Council and the Department of Science and Innovation (TdO).

## § Network for Genomic Surveillance in South Africa (NGS-SA)

Shareef Abrahams^1^, Luiz Carlos Junior Alcantara^2^, Arghavan Alisoltani-Dehkordi^3,4^, Mushal Allam^5^, Jinal N Bhiman^5,6^, Mary-Ann Davies^7,8^, Deelan Doolabh^9^, Susan Engelbrecht^10^, Vagner Fonseca^11^, Marta Giovanetti^2^, Allison J Glass^6,12^, Adam Godzik^4^, Dominique Goedhals^13^, Diana Hardie^14^, Marvin Hsiao^14^, Arash Iranzadeh^4^, Arshad Ismail^5^, Stephen Korsman^14^, Sergei L Kosakovsky Pond^15^, Oluwakemi Laguda-Akingba^1,16^, Jose Lourenco^17^, Gert Marais^14^, Darren Martin^9,18^, Caroline Maslo^19^, Koleka Mlisana^20,21^, Thabo Mohale^5^, Nokukhanya Msomi^22^, Innocent Mudau^9^, Francesco Petruccione^23,24^, Wolfgang Preiser^10^, Emmanuel James San^11^, Bryan Trevor Sewell^25^, Lynn Tyers^9^, Gert Van Zyl^10^, Anne von Gottberg^5,6^, Sibongile Walaza^5,26^, Steven Weaver^15^, Constantinos Kurt Wibmer^5^, Carolyn Williamson^9,14,21^, Denis York^27^.

^1^National Health Laboratory Service, Port Elizabeth, South Africa. ^2^Laboratorio de Flavivirus, Fundacao Oswaldo Cruz, Rio de Janeiro, Brazil. ^3^Division of Medical Virology, Department of Pathology, University of Cape Town, Cape Town, South Africa. ^4^Division of Biomedical Sciences, University of California Riverside School of Medicine, Riverside, California, USA. ^5^National Institute for Communicable Diseases of the National Health Laboratory Service, Johannesburg, South Africa. ^6^School of Pathology, Faculty of Health Sciences, University of the Witwatersrand, Johannesburg, South Africa. ^7^Centre for Infectious Disease Epidemiology and Research, University of Cape Town, Cape Town, South Africa. ^8^Western Cape Government: Health, Cape Town, South Africa. ^9^Division of Medical Virology, Institute of Infectious Disease and Molecular Medicine, University of Cape Town, Cape Town, South Africa. ^10^Division of Medical Virology at NHLS Tygerberg Hospital and Faculty of Medicine and Health Sciences, Stellenbosch University, Cape Town, South Africa. ^11^KwaZulu-Natal Research Innovation and Sequencing Platform (KRISP), Department of Laboratory Medicine and Medical Sciences, University of KwaZulu-Natal, Durban, South Africa. ^12^Department of Molecular Pathology, Lancet Laboratories, Johannesburg, South Africa. ^13^Division of Virology at NHLS Universitas Academic Laboratories, University of The Free State, Bloemfontein, South Africa. ^14^Division of Medical Virology at NHLS Groote Schuur Hospital, University of Cape Town, Cape Town, South Africa. ^15^Institute for Genomics and Evolutionary Medicine, Temple University, Philadelphia, Pennsylvania, USA. ^16^Department of Laboratory Medicine and Pathology, Faculty of Health Sciences, Walter Sisulu University, Mthatha, South Africa. ^17^Department of Zoology, University of Oxford, Oxford, United Kingdom. ^18^Computational Biology Division, Department of Integrative Biomedical Sciences, University of Cape Town, Cape Town, South Africa. ^19^Department of Quality Leadership, Netcare Hospitals, Johannesburg, South Africa. ^20^National Health Laboratory Service, Johannesburg, South Africa. ^21^Centre for the AIDS Programme of Research in South Africa (CAPRISA), Durban, South Africa. ^22^Discipline of Virology, University of KwaZulu-Natal, School of Laboratory Medicine and Medical Sciences and National Health Laboratory Service, Durban, South Africa. ^23^Centre for Quantum Technology, University of KwaZulu-Natal, Durban, South Africa ^24^National Institute for Theoretical Physics (NITheP), KwaZulu-Natal, South Africa. ^25^Structural Biology Research Unit, Department of Integrative Biomedical Sciences, University of Cape Town, Rondebosch, South Africa. ^26^School of Public Health, Faculty of Health Sciences, University of the Witwatersrand, Johannesburg, South Africa. ^27^Molecular Diagnostics Services, Durban, South Africa.

## § § COMMIT-KZN Team

Moherndran Archary^1^, Kaylesh J. Dullabh^2^, Philip Goulder^3,4^, Guy Harling^3,5^, Rohen Harrichandparsad^6^, Kobus Herbst^3,7^, Prakash Jeena^1^, Thandeka Khoza^3^, Nigel Klein^3,8^, Henrik Kløverpris^3,9,10^, Alasdair Leslie^3,9^, Rajhmun Madansein^2^, Mohlopheni Marakalala^3,9^, Matilda Mazibuko^3^, Mosa Moshabela^11^, Ntombifuthi Mthabela^3^, Kogie Naidoo^12^, Zaza Ndhlovu^3,13^, Thumbi Ndung’u^3,9,14,15^, Kennedy Nyamande^16^, Nesri Padayatchi^12^, Vinod Patel^17^, Theresa Smit^3^, Adrie Steyn^3,18^, Emily Wong^3,18^.

^1^Department of Paediatrics and Child Health, University of KwaZulu-Natal, Durban, South Africa. ^2^Department of Cardiothoracic Surgery, University of KwaZulu-Natal, Durban, South Africa. ^3^Africa Health Research Institute, Durban, South Africa. ^4^Department of Paediatrics, Oxford, UK. ^5^Institute for Global Health, University College London, UK. ^6^Department of Neurosurgery, University of KwaZuluNatal, Durban, South Africa. ^7^South African Population Research Infrastructure Network, Durban, South Africa. ^8^Institute of Child Health, University College London, UK. ^9^Division of Infection and Immunity, University College London, London, UK. ^10^Department of Immunology and Microbiology, University of Copenhagen, Copenhagen, Denmark. ^11^College of Health Sciences, University of KwaZulu-Natal, Durban, South Africa. ^12^Centre for the AIDS Programme of Research in South Africa, Durban, South Africa. ^13^Ragon Institute of MGH, MIT and Harvard, Boston, USA. ^14^HIV Pathogenesis Programme, The Doris Duke Medical Research Institute, University of KwaZulu-Natal, Durban, South Africa. ^15^Max Planck Institute for Infection Biology, Berlin, Germany. ^16^Department of Pulmonology and Critical Care, University of KwaZulu-Natal, Durban, South Africa. ^17^Department of Neurology, University of KwaZulu-Natal, Durban, South Africa. ^18^Division of Infectious Diseases, University of Alabama at Birmingham.

---

**Figure S1:**
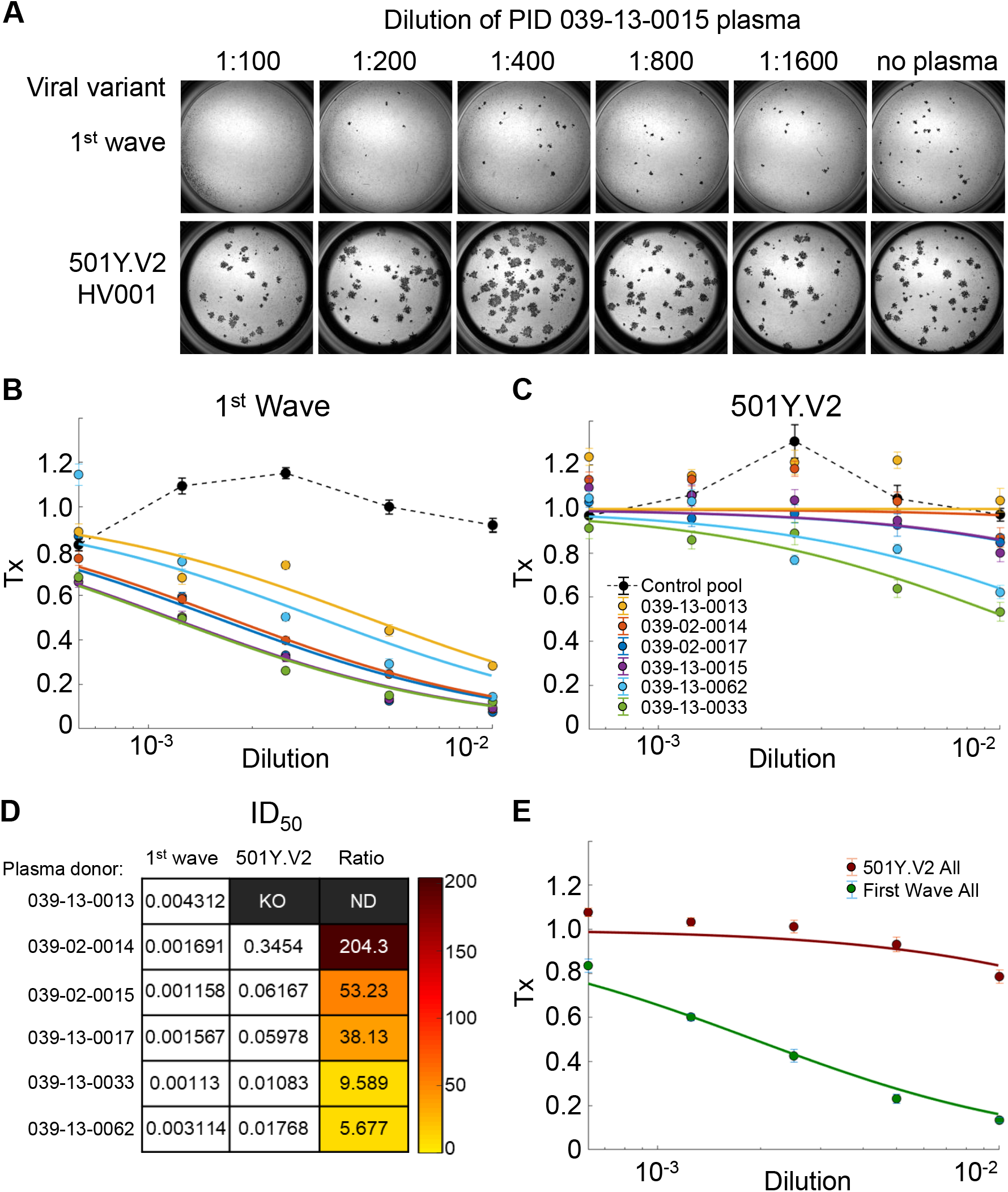
Neutralization of first wave and 501Y.V2 variants by convalescent plasma from first wave infections using equal infection incubation times. (A) A representative focus forming assay using plasma from participant 039-13-0015. Plasma neutralization of (B) first wave virus and (C) 501Y.V2 variants (501Y.V2.HV001 and 501Y.V2.HVdF002). Colored circles represent means and standard errors from 8 independent neutralization experiments using plasma from 6 convalescent participants who were infected by first wave variants in the first peak of the pandemic in South Africa. Correspondingly colored lines are fits of the sigmoidal equation with *ID*_50_ as the fitted parameter. Data from both 501Y.V2 variants was combined as separate experiments to obtain a more accurate fit of the data using a sigmoidal function since the declines in 501Y.V2 infection were small in the range of plasma concentrations used. The matched infections with first wave virus which were done in parallel with each 501Y.V2 variant were also combined. One experiment was removed in the process of quality control due to plate edge effects, which were subsequently corrected by adding sterile water between wells. Black points represent a pool of plasma from three uninfected controls. The transmission index (Tx) is the number of foci in the presence of the plasma dilution normalized by the number of foci in the absence of plasma. (D) Plasma *ID*_50_ values and ratios for first wave and 501Y.V2 variants. Knockout (KO) was scored as *ID*_50_ *>* 1. ND, not defined. (E) Mean and standard error across all plasma donors.

**Figure S2:**
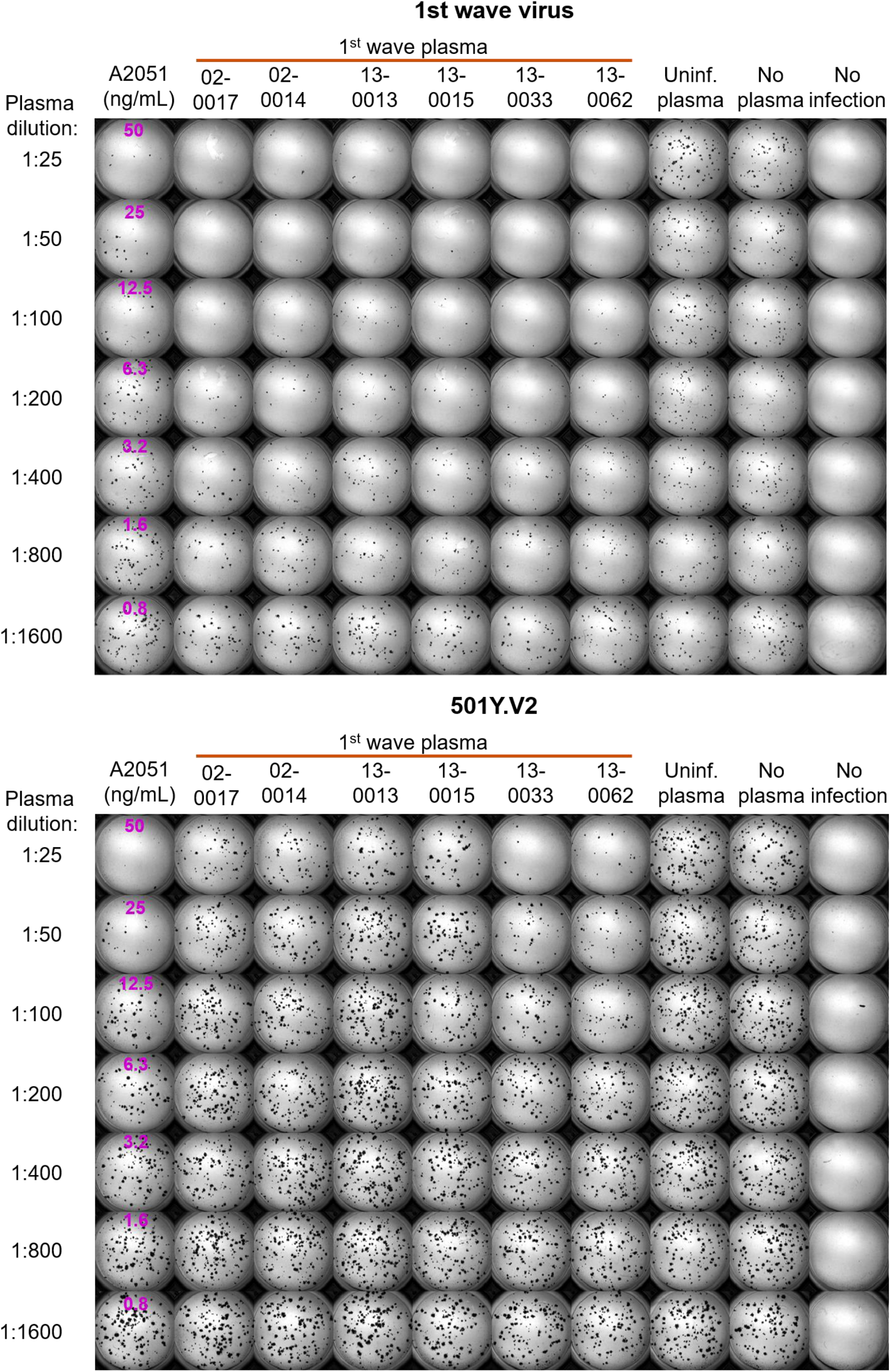
Neutralization of first wave and 501Y.V2 by convalescent plasma: Representative experiments of first set of participant plasma tested. Top montage shows neutralization of first wave virus, bottom montage shows neutralization of 501Y.V2. Rows are plasma dilutions, ranging from 1:25 to 1:1600. Last three columns are plasma from a pool of uninfected participants, the no plasma control, and no virus, respectively. First column is the A2051 NAb, with antibody concentrations in ng/mL (magenta). First wave plasma donors are marked with a red line, second wave plasma donors are marked with a blue line.

**Figure S3:**
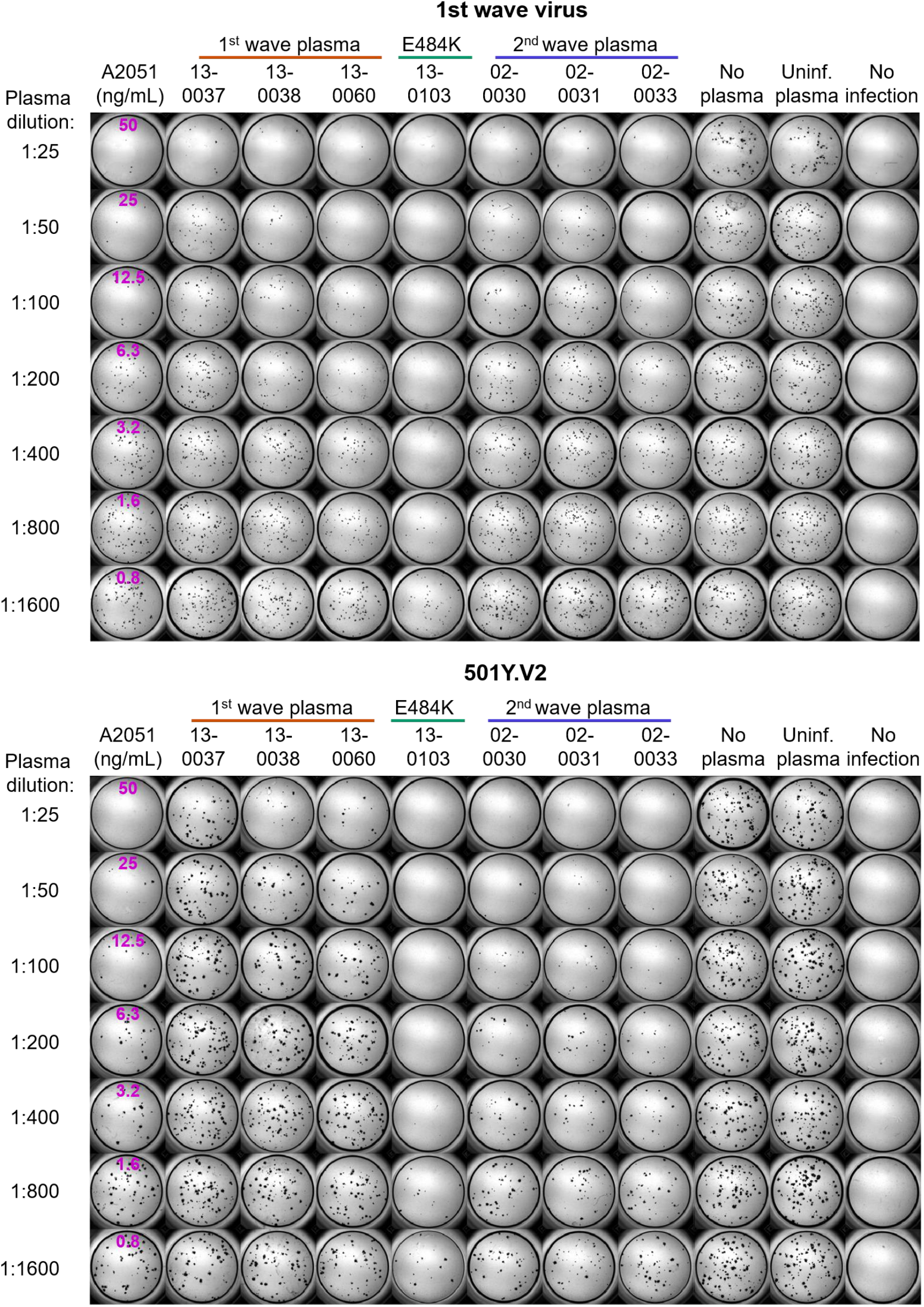
Neutralization of first wave and 501Y.V2 by convalescent plasma: Representative experiments of second set of participant plasma tested. Top montage shows neutralization of first wave virus, bottom montage shows neutralization of 501Y.V2. Rows are plasma dilutions, ranging from 1:25 to 1:1600. Last three columns are plasma from a pool of uninfected participants, the no plasma control, and no virus, respectively. First column is the A2051 NAb, with antibody concentrations in ng/mL (magenta). First wave plasma donors are marked with a red line, second wave plasma donors are marked with a blue line, the plasma donor showing the E484K mutation only is marked with a green line.

**Figure S4:**
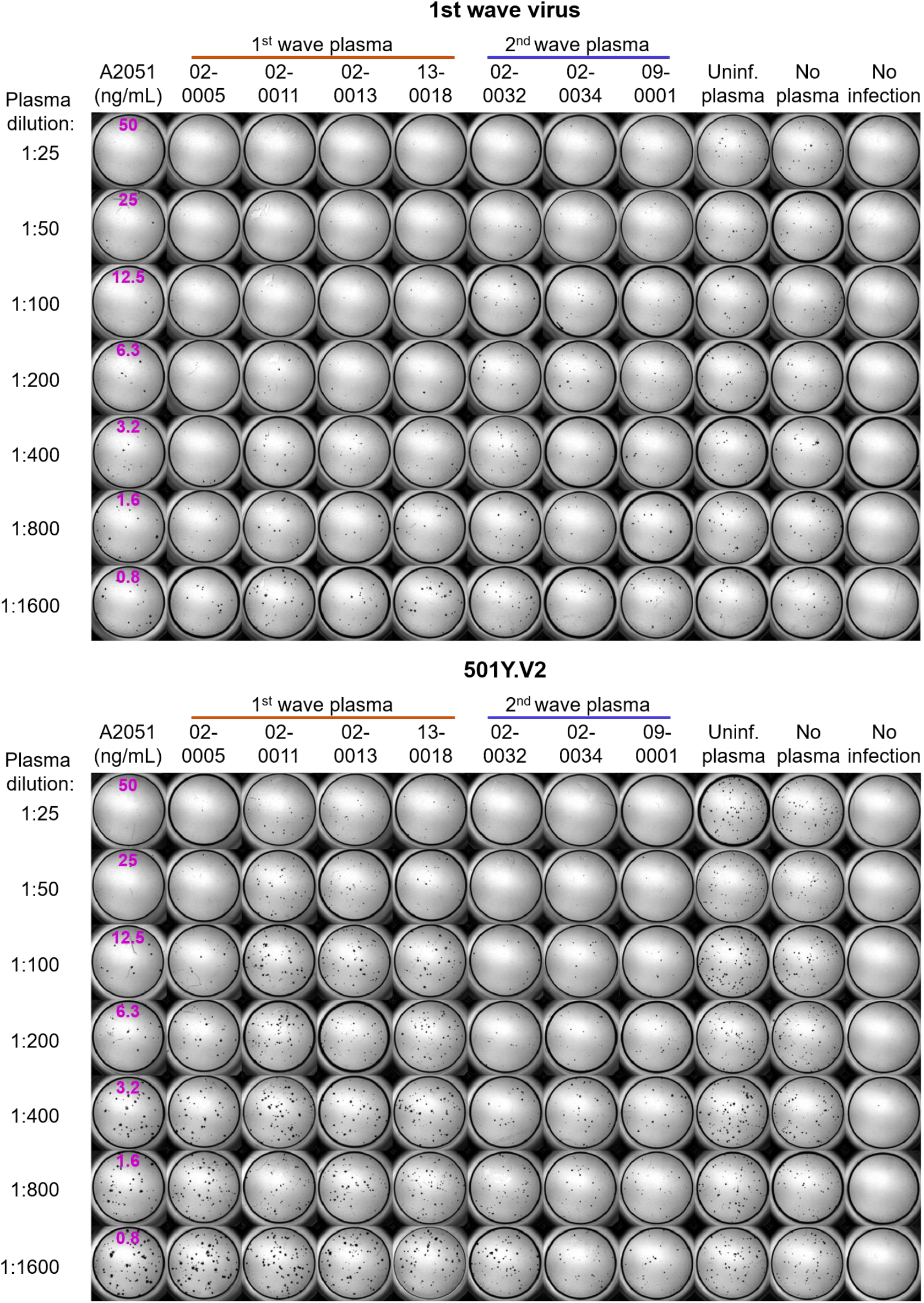
Neutralization of first wave and 501Y.V2 by convalescent plasma: Representative experiments of third set of participant plasma tested. Top montage shows neutralization of first wave virus, bottom montage shows neutralization of 501Y.V2. Rows are plasma dilutions, ranging from 1:25 to 1:1600. Last three columns are plasma from a pool of uninfected participants, the no plasma control, and no virus, respectively. First column is the A2051 NAb, with antibody concentrations in ng/mL (magenta). First wave plasma donors are marked with a red line, second wave plasma donors are marked with a blue line.

**Figure S5:**
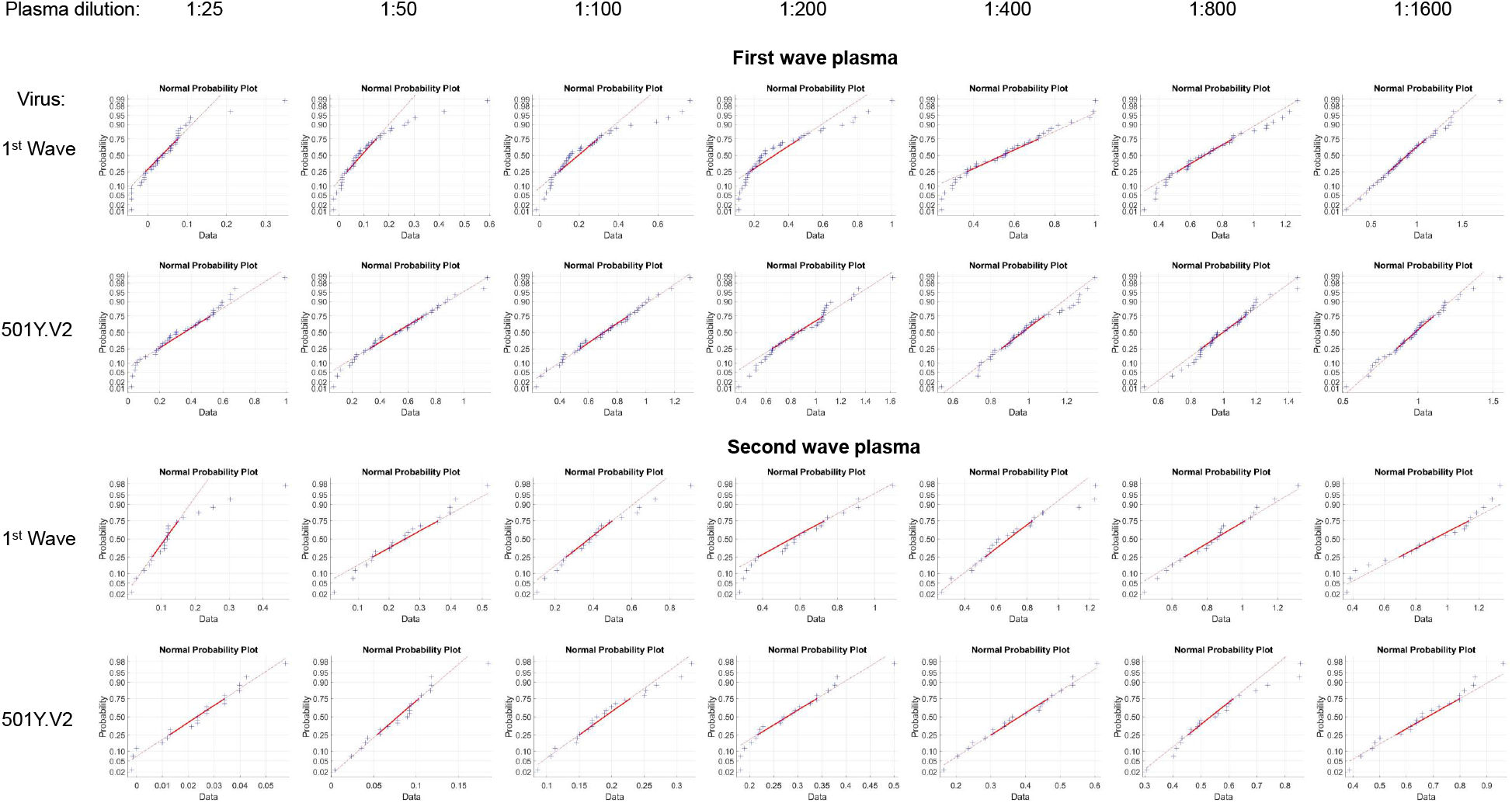
Fit of combined data for each plasma dilution to a normal distribution. The Matlab2019b function normplot was used to assess the fit of the data (blue crosses) to a normal distribution (solid red line). For each plot, one data point is the Tx result for one experiment for one participant at the specified dilution. Number of total experiments per viral variant was n=42 for first wave plasma, and n=21 for second wave plasma. Lack of pronounced curvature of the data in the range of the solid line indicates that a the data is a reasonably good fit to a normal distribution. see https://www.mathworks.com/help/stats/normplot.html for additional information.

**Table S1:** Plasma donor characteristics.

| Cohort ID | Sex | Age (yrs) | HIV status | HIV viral load (copies/mL) | Supplemental oxygen | Date of symptom onset | Days between symptom onset and plasma collection | Days between symptom onset and last positive qPCR |
| --- | --- | --- | --- | --- | --- | --- | --- | --- |
| 039-02-0005 | M | 50-59 | Negative | - | No | Mid-June | 29 | 8 |
| 039-13-0015 | F | 40-49 | Negative | - | No | Mid-June | 26 | 12 |
| 039-13-0033 | F | 30-39 | Negative | - | No | Late June | 30 | 23 |
| 039-02-0013 | F | 70+ | Negative | - | Yes | Late June | 29 | 15 |
| 039-13-0013 | F | 50-59 | Positive | <40 | No | Late June | 30 | 10 |
| 039-13-0018 | F | 40-49 | Negative | - | No | Late June | 28 | 14 |
| 039-02-0014 | F | 60-69 | Negative | - | No | Early July | 27 | 20 |
| 039-02-0011 | F | 40-49 | Positive | <40 | No | Early July | 32 | 32 |
| 039-13-0060 | M | 40-49 | Positive | <40 | No | Mid-July | 38 | 31 |
| 039-02-0017 | F | 60-69 | Negative | - | Yes | Mid-July | 28 | 7 |
| 039-13-0062 | M | 60-69 | Negative | - | No | Early August | 26 | 12 |
| 039-13-0103 | M | 60-69 | Negative | - | Yes | Late September | 37 | 17 |
| 039-13-0037 | M | 30-39 | Positive | <40 | No | Asymptomatic | 29* | 8† |
| 039-13-0038 | M | 30-39 | Negative | - | No | Asymptomatic | 29* | 8† |
| 039-02-0031 | F | 40-49 | Negative | - | Yes | Late December | 34 | 13 |
| 039-02-0032 | M | 40-49 | Positive | <40 | Yes | Late December | 43 | 12 |
| 039-02-0033 | M | 50-59 | Negative | - | No | Late December | 35 | 42 |
| 039-02-0030 | F | 40-49 | Positive | <40 | Yes | Late December | 30 | 9 |
| 039-02-0034 | F | 30-39 | Negative | - | Yes | Late December | 32 | 5‡ |
| 039-09-0001 | F | 60-69 | Negative | - | Yes | Early January | 29 | 29 |

\*Asymptomatic cases; plasma collected 29 days after positive diagnostic swab for these two participants
†Last positive qPCR collected 8 days after diagnostic swab collection for two participants
‡Only single qPCR test positive at diagnosis

**Table S2:**
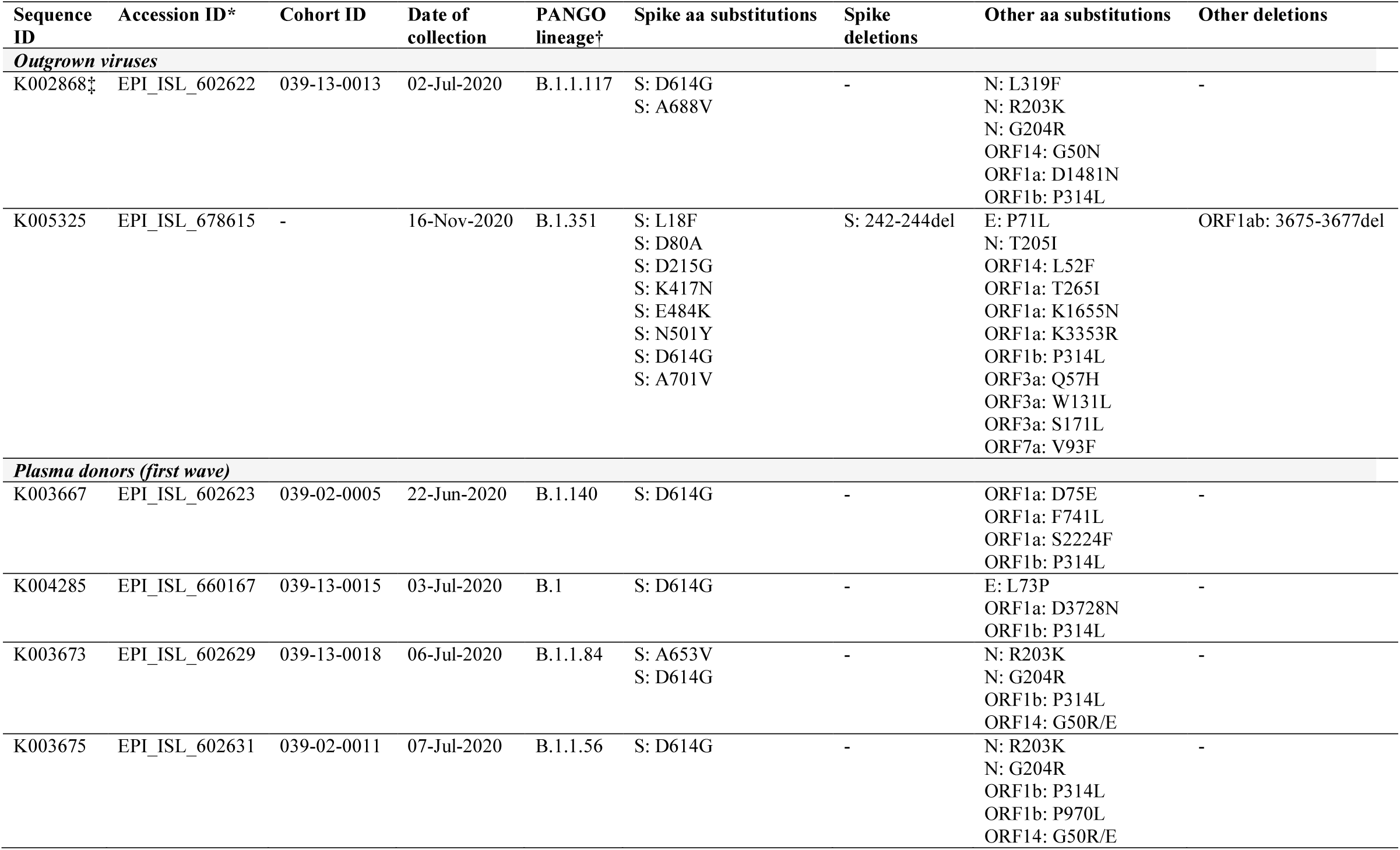
Mutation profile for the genomes of the outgrown viruses and for the infecting viruses of convalescent plasma donors.

**Table S3:** Mutation profile for the genomes of the outgrown viruses and for the infecting viruses of convalescent plasma donors continued.

|  |  |  |  |  |  |  |  |  |
| --- | --- | --- | --- | --- | --- | --- | --- | --- |
| K003668 | EPI_ISL_602624 | 039-02-0013 | 10-Jul-2020 | B.1.1.117 | S: A688V<br>S: D614G | - | N: L139F<br>N: R203K<br>N: G204R<br>ORF1b: P314L<br>ORF14: G50R/E | - |
| K004289 | EPI_ISL_660170 | 039-02-0014 | 14-Jul-2020 | B.1.1.56 | S: D614G | - | E: L73P<br>N: R203K<br>N: G204R<br>ORF14: G50N<br>ORF1b: P314L<br>ORF1b: T1522I | - |
| K004293 | EPI_ISL_660174 | 039-13-0038 | 16-Jul-2020 | H.1 | S: D614G<br>S: Q677H | - | N: R203K<br>N: G204R<br>ORF1a: P1497L<br>ORF1a: D3728N<br>ORF1b: P314L<br>ORF14: G50N | - |
| K004291 | EPI_ISL_660172 | 039-13-0033 | 17-Jul-2020 | B.1 | S: D614G | - | N: T148A<br>ORF10: A28V<br>ORF1a: K2511R<br>ORF1a: V3858I<br>ORF1b: P314L | - |
| K004292 | EPI_ISL_660173 | 039-13-0037 | 24-Jul-2020 | B.1.1.117 | S: D614G<br>S: A688V<br>S: M731I | - | N: L139F<br>N: R203K<br>N: G204R<br>ORF3a: Q245K<br>ORF1b: P314L | - |
| K004295 | EPI_ISL_660176 | 039-02-0017 | 28-Jul-2020 | B.1.140 | S: D614G | - | ORF1a: F1178S<br>ORF1b: P314L | - |
| K004300 | EPI_ISL_660180 | 039-13-0060 | 17-Aug-2020 | B.1.1.56 | S: D614G | - | E: L73P<br>N: R203K<br>N: G204R<br>ORF1a: R2267G<br>ORF1b: P314L<br>ORF1b: A2589T<br>ORF14: G50N | - |
| K004302 | EPI_ISL_660181 | 039-13-0062 | 18-Aug-2020 | B.1.1.1 | S: D614G | - | N: R203K<br>N: G204R | - |

**Table S4:**
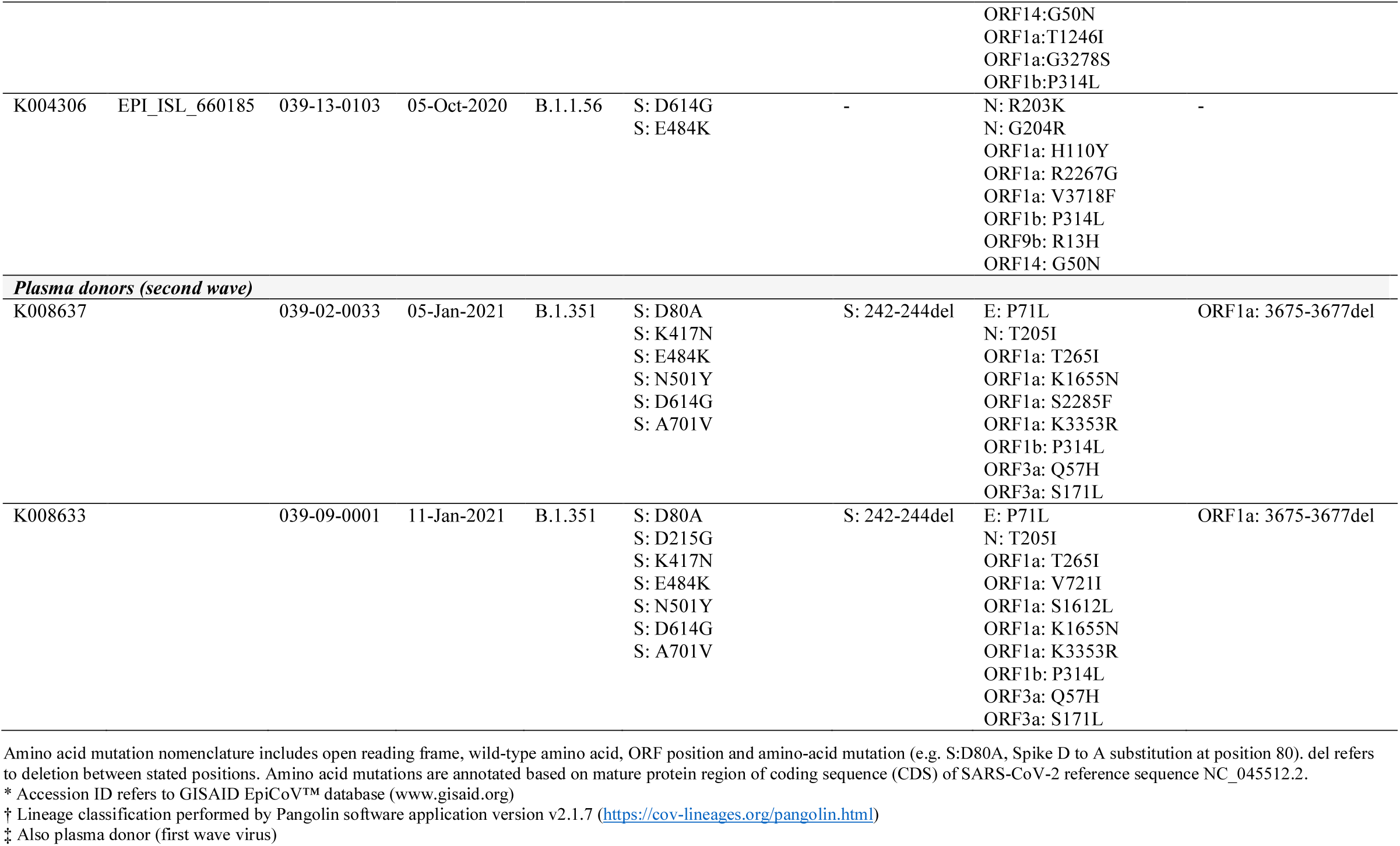
Mutation profile for the genomes of the outgrown viruses and for the infecting viruses of convalescent plasma donors continued.

